# Modeling the impact of adherence to U.S. isolation and masking guidance on SARS-CoV-2 transmission in office workplaces in 2021-2022

**DOI:** 10.64898/2026.04.14.26350639

**Authors:** Maria Garcia Quesada, Karina Wallrafen-Sam, Moses Chapa, Faruque Ahmed, Obianuju Genevieve Aguolu, Noureen Ahmed, Saad B. Omer, Benjamin A. Lopman, Samuel M. Jenness

## Abstract

Non-pharmaceutical interventions (NPIs) have been important for controlling SARS-CoV-2 transmission, particularly before and during initial vaccine rollout. During the pandemic, the US Centers for Disease Control and Prevention issued isolation and masking guidance in case of COVID-19-like illness, a positive SARS-CoV-2 test, or known exposure to SARS-CoV-2. However, the impact of this guidance on mitigating transmission in office workplaces is unclear. We used a network-based mathematical model to estimate the impact of this guidance on SARS-CoV-2 transmission among office workers and their communities. The model represented social contacts in the home, office, and community. We used data from the CorporateMix study to parametrize social contacts among office workers and calibrated the model to represent the COVID-19 epidemic in Georgia, USA from January 2021 through August 2022. In the reference scenario (58% adherence to guidance among office workers and the broader population), workplace transmission accounted for a small fraction of total infections. Reducing adherence among office workers to 0% increased workplace transmissions by 27.1% and increasing adherence to 75% reduced workplace transmission by 7.0%. Increasing adherence to 75% among office workers had minimal impact on symptomatic cases and deaths; increasing it among the broader population was more effective in reducing office worker cases and deaths. In our model, moderate adherence to recommended NPIs in workplaces was effective in reducing transmission, but increasing adherence had limited benefit given workplaces that have low contact intensity and hybrid work arrangements. These results underscore the public health benefits of community-wide adoption of recommended NPIs.

## Background

The COVID-19 pandemic resulted in devastating morbidity and mortality in the United States, where both reported incidence and mortality rates have been among the highest in the world.^1^ As of February 2026, COVID-19 has caused over 1.2 million deaths in the US.^2^ The pandemic underscored the critical need for effective and adaptable public health guidance, especially as companies navigated decisions around when and how employees could return to physical office spaces. To address this need, the US Centers for Disease Control and Prevention (CDC) issued comprehensive guidance aimed at minimizing SARS-CoV-2 transmission. Recommendations evolved over the course of the pandemic, generally going from more to less stringent following the widespread introduction of vaccines.^3^ After vaccines became widely available, CDC guidance focused on isolation for symptomatic and SARS-CoV-2 infected individuals, and masking and testing following exposure to the virus.^4,5^

Although isolation and masking have been associated with reducing SARS-CoV-2 transmission in the community, the potential impact of these interventions in non-healthcare office spaces is unclear.^6,7^ Mathematical modeling provides a framework to simulate how isolation and masking may mitigate transmission, and quantify the resulting impact on COVID-19 cases. However, these models require an understanding of contact patterns across age groups and in relevant settings, along with how those contact patterns change in response to public health guidance. To this end, a serial cross-sectional study (Corporate Mix) measured social contact patterns among employees from five companies based in Atlanta, Georgia, USA. Surveys were administered at multiple time points during April 2020–June 2022, a time period during which work from home and hybrid work arrangements had become commonplace.^8^ The study aimed to collect detailed data on the contacts of office workers both in and out of the office, so these could be incorporated into mathematical models to simulate the potential impact of different intervention strategies.

In this study, we use a mathematical model of SARS-CoV-2 transmission that can represent these complex, multi-layered contact networks to evaluate the potential impact of CDC guidance for isolation and masking on COVID-19 among non-healthcare office workers in Georgia, USA in the post-vaccine period. Specifically, the study examines whether improved adherence to guidance among office workers and among the total population may have been an effective strategy to reduce cases, considering relatively low contact rates in office settings resulting from a shift towards hybrid work (i.e., optional work-from-home arrangements).

## Methods

A SARS-CoV-2 network-based mathematical model was used to simulate vaccination rates and COVID-19 cases in Georgia, USA from January 2021 through August 2022. This period encapsulates the time between when COVID-19 vaccines first became available in Georgia through the start of the bivalent booster vaccine rollout. The model was developed using EpiModelCOVID, an extension of the EpiModel software platform, which uses the statistical framework of exponential random graph models (ERGMs) to estimate and simulate from statistical models dynamic contact networks and has been extensively used and validated.^9^ We modeled varying adherence to CDC isolation and masking guidance as reductions in contact intensity (i.e., how much persons reduced their contacts while isolating) and transmission probabilities per contact, respectively. The model code and software are available on GitHub (https://github.com/EpiModel/COVID-Corporate).

### Model Structure

The model represented 100,000 persons (agents) of all ages who were each assigned an initial age according to Georgia’s 2020 age pyramid.^10^ New agents could enter the model through birth and existing agents could exit the model through death, all-cause or COVID-19-specific. A subset of agents ages 20-89 years old were categorized as office workers based on 1) the proportion of U.S. adults who were employed by age group in 2022, and 2) the proportion of those employed who worked in an office setting (e.g., 38.38% of 30-49 year olds were office workers; Table S1).^11^ All agents were members of two contact network layers representing the household and the community, and office worker agents were additionally members of a third contact network layer representing the workplace, where they only had contacts with other office workers. Contacts across all layers represented those longer than 15 minutes given these are thought to be most relevant for SARS-CoV-2 transmission.^12^

Contacts (edges) in the household network layer were persistent, meaning that they remained fixed across the simulation period. All agents were assigned to a household according to an algorithm as previously described,^13^ such that household structures by size and age group resembled those in the U.S. Census. Household edges were specified across all individuals assigned to each household, and each edge was limited to a single household.

Edges in the workplace layer were also persistent. Contact patterns in the workplace were estimated based on workplace contacts longer than 15 minutes reported in the round of the Corporate Mix study conducted during March–June 2022.^8^ Average total daily contacts in the workplace were 0.72, and we parameterized total daily contacts by age group for 20-69 year olds and by within age group mixing for all age groups (Table 1).

**Table 1.**
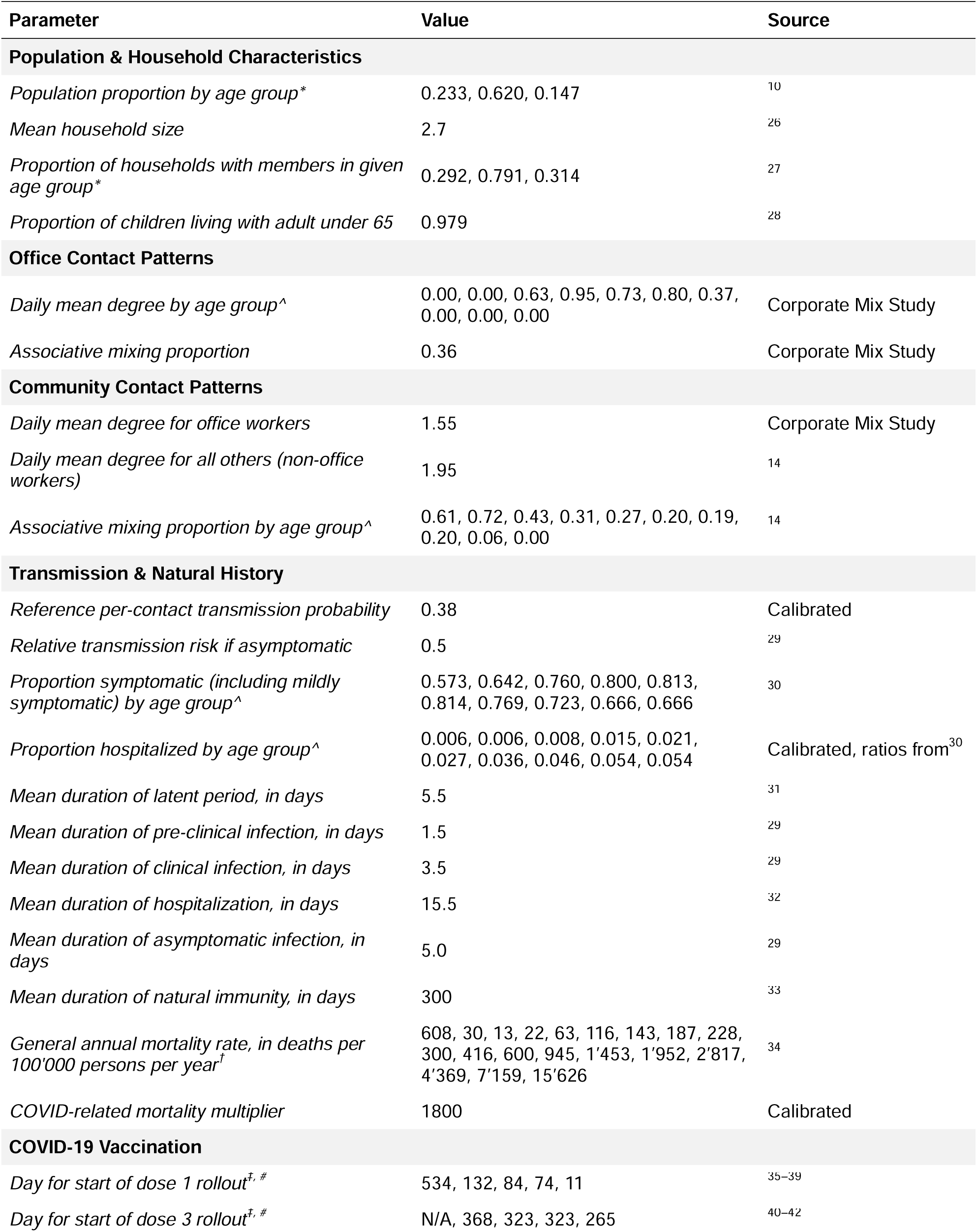
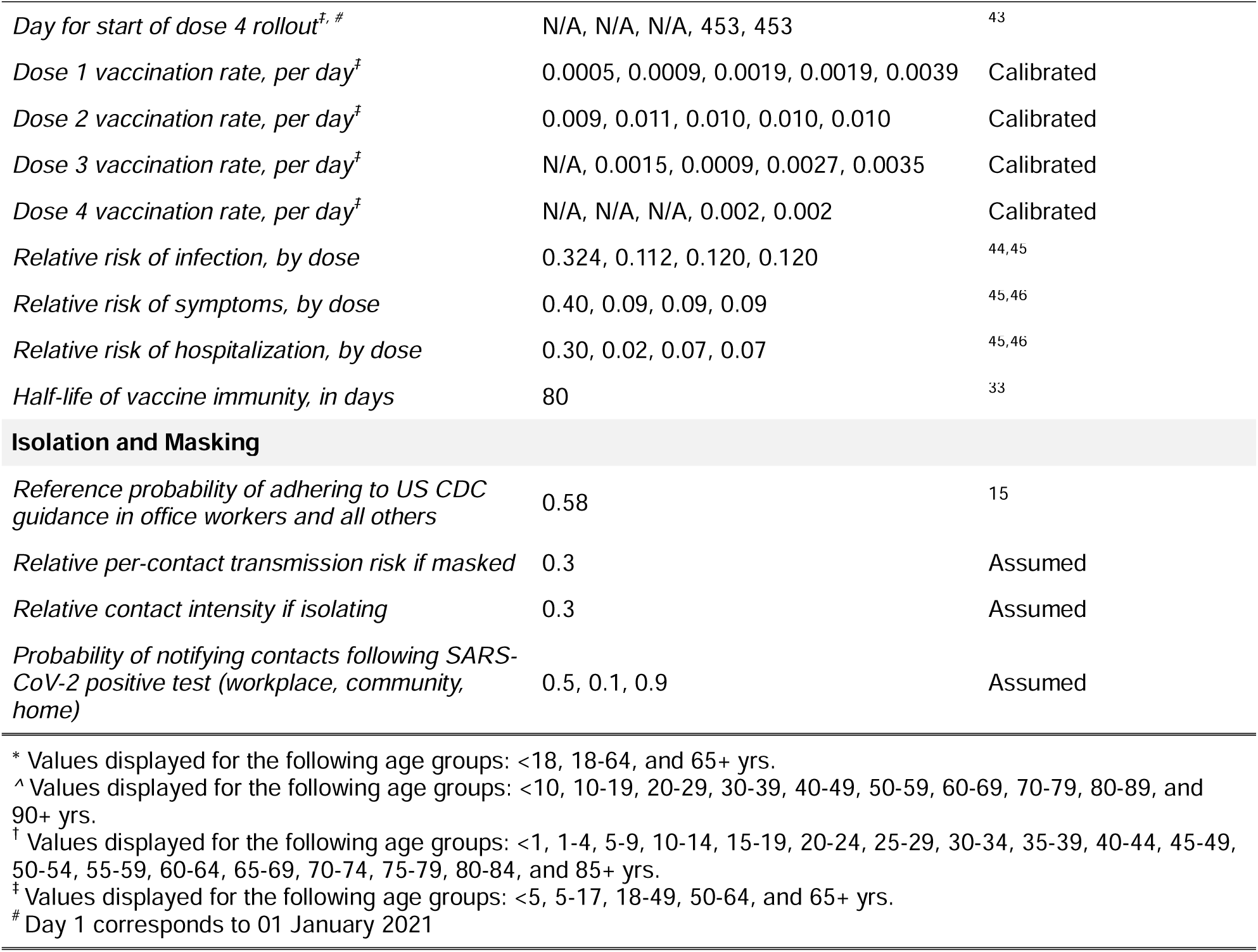
Parameters for SARS-CoV-2 network transmission model representing the COVID-19 pandemic in Georgia, USA, between January 2021 and August 2022.

Edges in the community layer were dynamic and non-persistent, meaning that they were re-simulated based on an ERGM at each modeled time step; each time step represented one day. The community layer represented all non-household and non-office contacts for office workers, and all non-household contacts for others (i.e., including other work, school, etc.). For office workers, average total daily contacts in the community were 1.55, as reported in the Corporate Mix data. For others, average total daily contacts in the community were 1.95 based on the POLYMOD social mixing study.^14^ For 0-69 year olds, within age group mixing was also parametrized based on POLYMOD data (Table 1).^14^

The model used a SEIRS disease framework (i.e., susceptible, exposed, infected, recovered, susceptible) to represent the natural history of COVID-19, with transitions occurring stochastically between disease states. Following a contact between a susceptible and an infected agent, the probability that the susceptible agent became infected depended on their vaccination status, whether the infected agent was symptomatic, and whether the contact occurred in the household layer. If infected, agents could become symptomatic or remain asymptomatic, and those symptomatic could become hospitalized. The probabilities of symptoms and hospitalization varied by age and vaccination status, and age-specific mortality rates were higher for those hospitalized. Agents could also become vaccinated at age-specific rates given they were not currently symptomatic, had not tested positive in the last 10 days, and were eligible for vaccination based on their age group. Vaccination reduced the risk of infection, developing symptoms, and becoming hospitalized. Both natural and vaccine immunity waned over time. Key model parameters are provided in Table 1.

### Modeling CDC Isolation and Masking Guidance

Agents in the model could follow different CDC isolation and masking guidance in response to each of three triggers: experiencing COVID-19-like illness (symptomatic infection), testing positive for SARS-CoV-2, or being notified of an exposure to a recent contact who tested positive for SARS-CoV-2.^4,5^

If an agent developed COVID-19-like illness (i.e., symptoms) and was determined to adhere to the guidance, they isolated for at least five days, with day zero being the day of symptom onset, or until symptoms subsided, whichever came later. If an agent became hospitalized, they isolated for at least 10 days or until symptoms subsided. Regardless of hospitalization, agents masked through day 10 or until they tested negative.

If an agent tested positive for SARS-CoV-2 while asymptomatic and was determined to adhere to the guidance, they isolated for at least five days, with day 0 being the day they were tested. If they did not develop symptoms, they ended isolation after day five. If they developed symptoms, the isolation ‘clock’ reset to zero on the day of symptom onset and they followed the guidance for COVID-19-like illness described above. Regardless of symptoms, agents masked through day 10 or until they tested negative.

If an agent was notified of a SARS-CoV-2 exposure and was determined to adhere to the guidance, they wore a mask through day 10 following the exposure, with day zero being the day of the (assumed) exposure, and got tested on day six. If they tested negative, they continued to wear a mask through day 10. If they tested positive, they began isolating and followed the guidance for a positive SARS-CoV-2 test. If they developed symptoms, they followed the guidance for COVID-19-like illness.

Whether an agent adhered to the CDC guidance following a trigger was represented stochastically by a guidance adherence probability parameter. The adherence parameter in reference scenarios, for both office workers and all others, was based on a survey that found 58% adherence to masking in Georgia residents in September 2020; this is consistent with other studies of adherence to disease-mitigation guidance.^15,16^ A separate notification probability parameter, specific to each of the network layers, determined whether an agent who tested positive for SARS-CoV-2 notified their recent contacts in each of the layers. The reference scenario parameter was assumed to be 90% for household contacts, 50% for office contacts, and 10% for community contacts. Isolation was modeled as a 70% reduction in contact intensity, while masking was modeled as a 70% reduction in infection probability per contact.

Testing occurred among all those adhering to guidance and either masking post-isolation or after five days of masking post-exposure. Routine disease screening also occurred stochastically among all agents regardless of adherence status; those who were symptomatic had a greater probability of testing than those who were non-symptomatically infected or not infected (Table 1). Tests were assumed to have a sensitivity of 80%.

### Model Calibration

Calibration was completed using an iterative rejection algorithm, by exploring ranges of values across the parameter space for uncertain parameters until best fit to target statistics was achieved by statistical and visual assessment. First, age- and dose-specific vaccination rates were calibrated to match vaccine coverage by age, dose, and month in Georgia.^17^ Second, the infection probability per contact was calibrated so that, after scaling for population size and adjusting for underreporting,^18^ simulated incident infections per month matched the confirmed infection counts reported by the Georgia Department of Public Health (GDPH).^19^ Modifiers for the infection probability per contact were also calibrated to boost and suppress cases at different time periods; this improved the fit to the observed variability in case rates resulting from SARS-CoV-2 variants and changes in population immunity and behaviors. Third, age-specific hospitalization proportions and a COVID-19 mortality multiplier were calibrated to match monthly COVID-19 related hospitalizations and deaths in Georgia.^19^

### Intervention Scenarios

Our calibrated model was considered the reference scenario, to which we compared the impact of interventions on parameters related to adherence to the CDC guidance on isolation and masking. These parameters included: overall adherence to guidance (reference: 58%), probability of notifying work contacts following a positive SARS-CoV-2 test (reference: 50%), and relative reduction in contact intensity during isolation (reference: 70%). Interventions tested both increases and decreases to relevant parameters, to examine both 1) the potential impact of improvements in adherence to CDC guidance beyond what was practiced (i.e., reference scenario), and 2) what may have been the impact of adherence to CDC guidance as practiced on disease outcomes. The primary intervention scenarios included no adherence to guidance (0%), worse adherence (25%), improved adherence (75%), and perfect adherence (100%). We tested the impact of varying these parameters among the total population, and among office workers and other agents separately while the non-intervention group remained at the reference value of 58%.

We also conducted two-dimensional sensitivity analyses by varying two intervention-related parameters across the parameter space (range: 0-1) and evaluated the impact on three outcomes: total persons entering isolation, infection rate, and mortality rate. The parameter combinations we evaluated included 1) adherence probability among office workers vs adherence probability among all others, 2) adherence probability among office workers vs notification probability at work, 3) adherence probability among office workers vs relative contact intensity, and 4) adherence probability among the total population vs relative contact intensity. Outcome values were visualized as contour plots to characterize how each combination of adherence probabilities jointly influences model projections.

### Model Outcomes

Outcomes from each scenario included the rate of agents entering isolation following a positive SARS-CoV-2 test or symptom onset, rate of agents beginning masking following a known exposure, incidence rates of SARS-CoV-2 infections, COVID-19 cases (i.e., symptomatic infections), and COVID-19 deaths. To examine how interventions impacted transmission in the workplace, we differentiated infections that occurred in the workplace layer of the network from those that occurred at home or in the community. To examine the impact of interventions on office workers, cases and deaths were tracked for the total population as well as for the subset of agents designated as office workers. We report the median and 50% simulation interval (SI) of each output across 250 simulations per scenario.

## Results

Our calibrated model in the reference scenario (58% adherence to guidance among office workers and others) achieved a reasonable fit to target statistics representing the COVID-19 epidemic in Georgia, USA during January 2021–August 2022. Figure 1 shows the fit of simulated monthly COVID-19 infections, hospitalizations, and deaths, while Figure 2 shows the fit of the simulated vaccine coverage by dose and age group.

**Figure 1.**
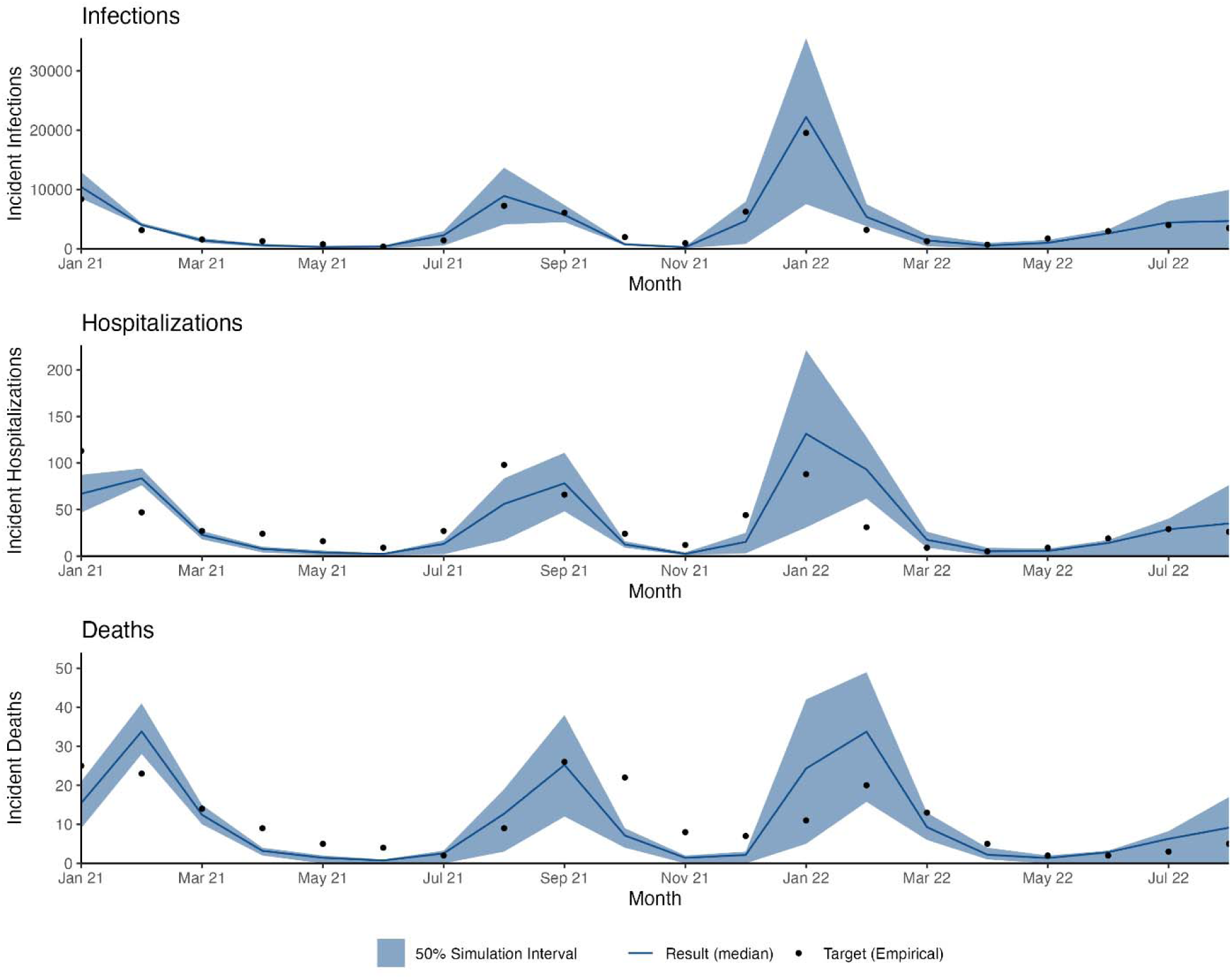
Modeled incident infections, hospitalizations, and deaths from a network transmission SARS-CoV-2 model compared to target statistics from Georgia, USA, from January 2021 through August 2022 for model calibration. Target incident infections, hospitalizations, and deaths estimated from Georgia Department of Public Health (GDPH) COVID-19 data.

**Figure 2.**
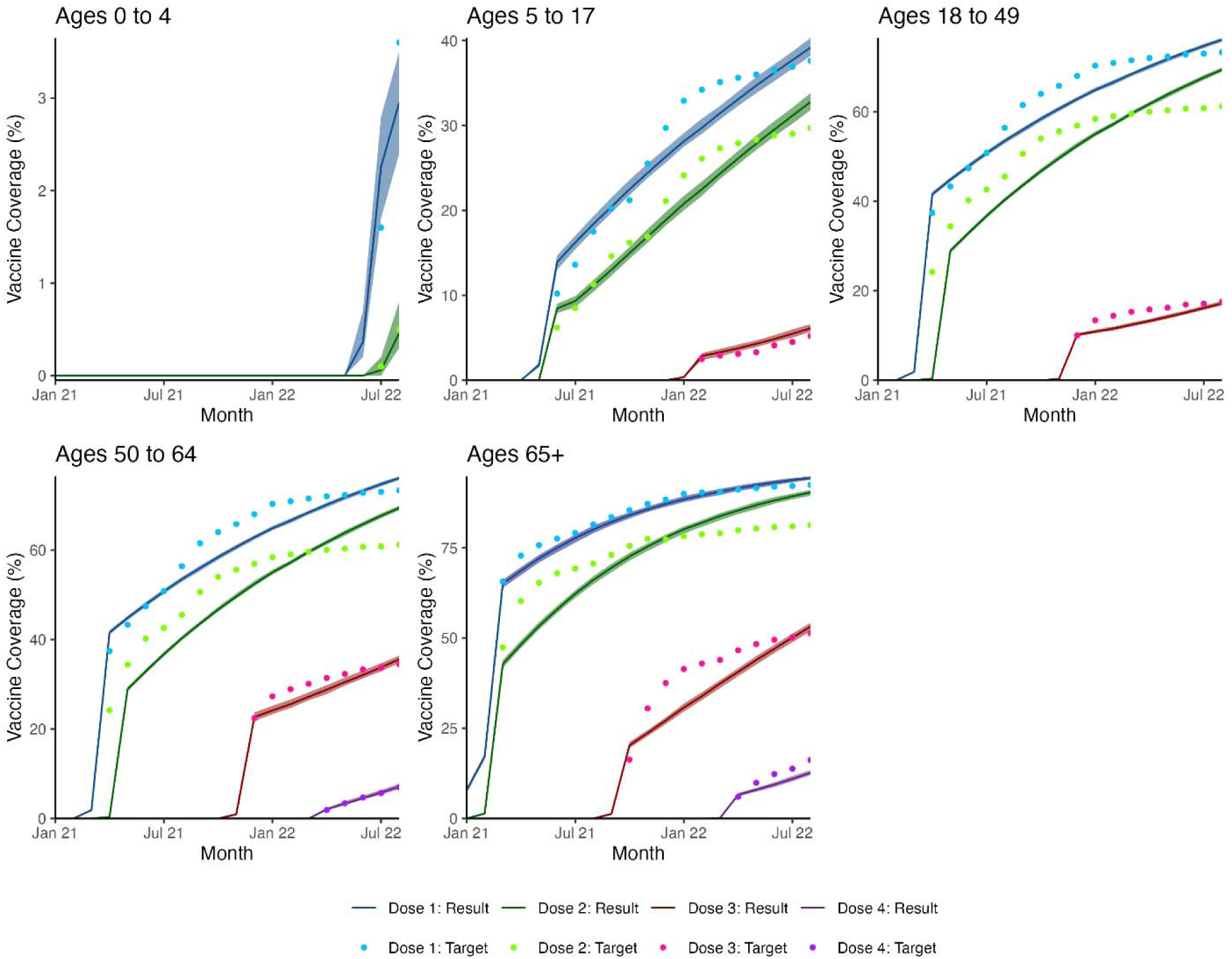
Modeled age- and dose-specific COVID-19 vaccination coverage from a network transmission SARS-CoV-2 model compared to target statistics from Georgia, USA, from January 2021 through August 2022 for model calibration. Target vaccination coverage estimated from US CDC COVID-19 vaccination rates in Georgia.

In the reference scenario, there were 131.8 (50% SI: 128.9, 139.0) infections, 95.5 (50% SI: 92.1, 98.1) cases, and 0.34 (50% SI: 0.32, 0.36) deaths per 100,000 person-days in the total population (Tables S2-S4). About 12.3% of all transmissions occurred in the workplace, or 16.2 (50% SI: 15.6, 16.8) infections per 100,000 person-days (i.e., excluding infections among office workers where transmission occurred at home or in the community, and infections among all others). Regardless of where transmission occurred, office workers experienced 101.6 (50% SI: 97.3, 104.6) cases and 0.39 (50% SI: 0.34, 0.42) deaths per 100,000 person-days (Table S3-S4).

Varying adherence to CDC guidance among office workers had meaningful effects on workplace transmission specifically. Compared to the reference scenario (58% adherence among both office workers and all others), reducing adherence to 0% among office workers and keeping it at 58% for all others, resulted in an additional 27.1% (50% SI: 23.7, 32.0) SARS-CoV-2 transmissions in the workplace (Figure 3). Improved adherence (75%) among office workers prevented an additional 7% (50% SI: 1.9, 12.9) of workplace transmissions compared to the reference scenario (Figure 3). However, this effect was limited to workplace transmissions; varying adherence among office workers had negligible impact on infections across the total population (Figure 3, Figure S1).

**Figure 3.**
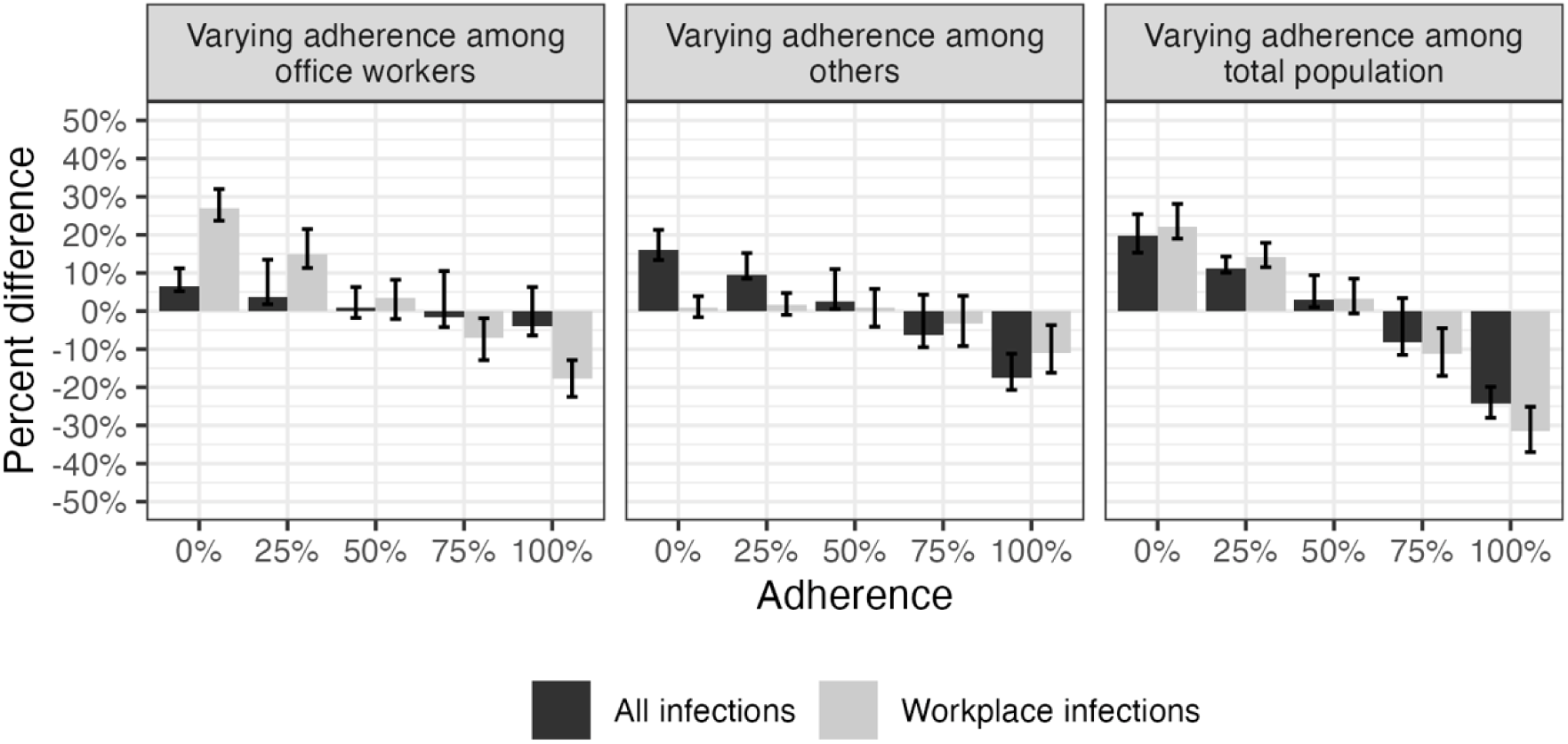
Percent change in total and workplace infection rates when varying adherence to U.S. isolation and masking guidance among office workers, others, and the total population in a SARS-CoV-2 network transmission model representing Georgia, USA, from January 2021 through August 2022. Infection rates measured as number of infections per 100,000 person-days. Adherence among non-intervention group is fixed at reference value (58%). For example, when varying adherence among office workers, adherence among all others is 58%. Workplace infections refer to infections among office workers where transmission occurred in the workplace (i.e., excludes infections among office workers where transmission occurred at home or in the community). The data are shown in tabular format in Table S2.

Changes in office worker adherence to CDC guidance had relatively small effects on symptomatic cases and deaths. No adherence (0%) among office workers resulted in 9.1% (50% SI: 6.9, 11.3) more office worker cases and 5.9% (50% SI: -5.7, 15.6%) more office worker deaths compared to the reference scenario (Tables S3-S4, Figure 4). Improved adherence (75%) among office workers only prevented an additional 2.0% (50% SI: -2.7, 7.1) of office worker cases and 2.0% (50% SI: -7.7, 13.5) of office worker deaths (Tables S3-S4, Figure 4). The impact of office worker adherence on total population cases and deaths was even smaller (Figure 4). Although peaks in cases were similar across levels of adherence over the course of the simulation period, greater adherence resulted in a greater proportion of cases isolating (Figure 5, Figure 6).

**Figure 4.**
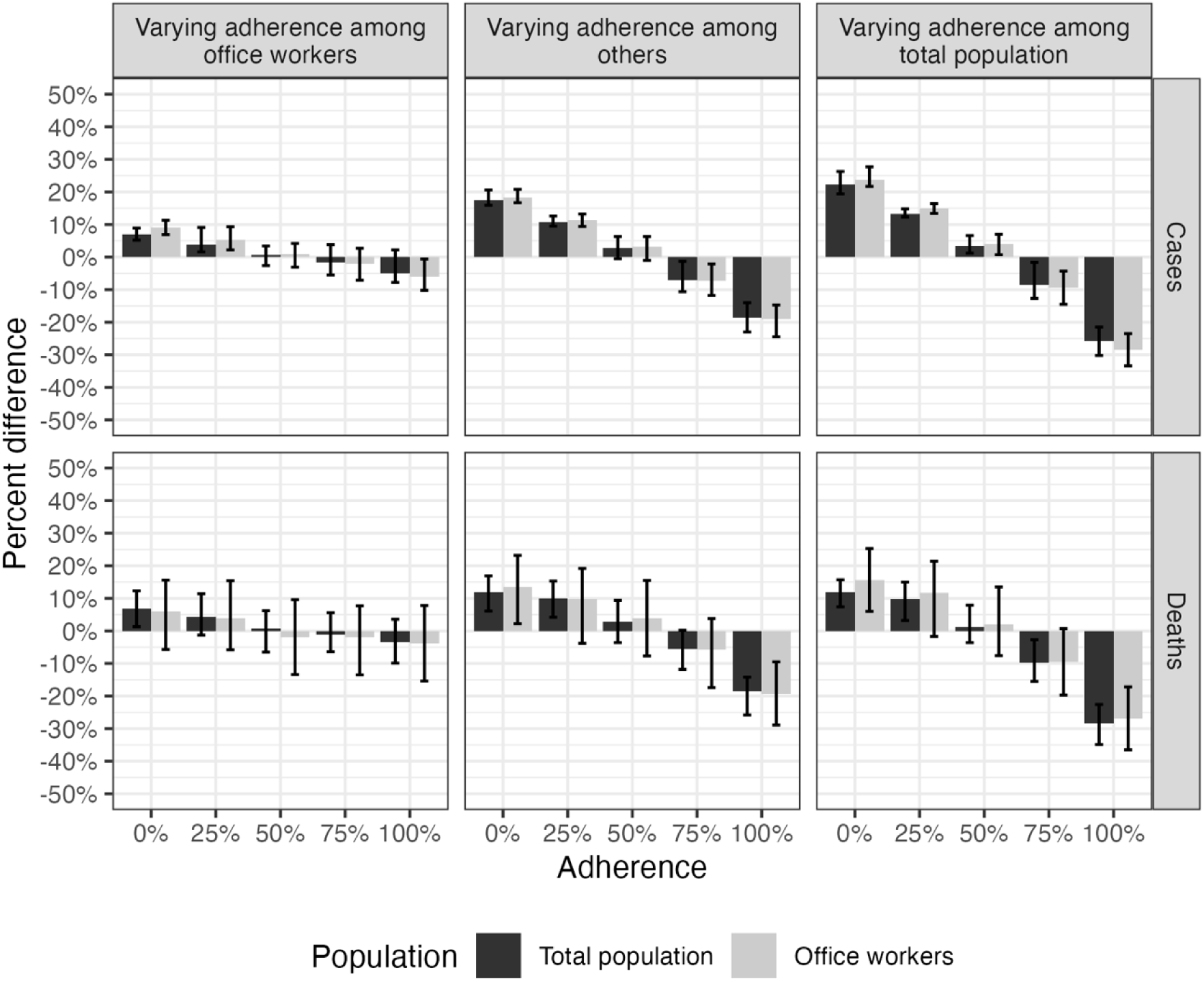
Percent change in case and death rates among the total population and office workers when varying adherence to U.S. isolation and masking guidance among office workers, others, and the total population in a SARS-CoV-2 network transmission model representing Georgia, USA, from January 2021 through August 2022. Case and death rates measured as number of cases or deaths per 100,000 person-days. Cases and deaths among office workers are inclusive of those where transmission occurred in the workplace as well as at home and in the community. Adherence among non-intervention group is fixed at reference value (58%). For example, when varying adherence among office workers, adherence among all others is 58%. The data are shown in tabular format in Tables S3 and S4.

**Figure 5.**
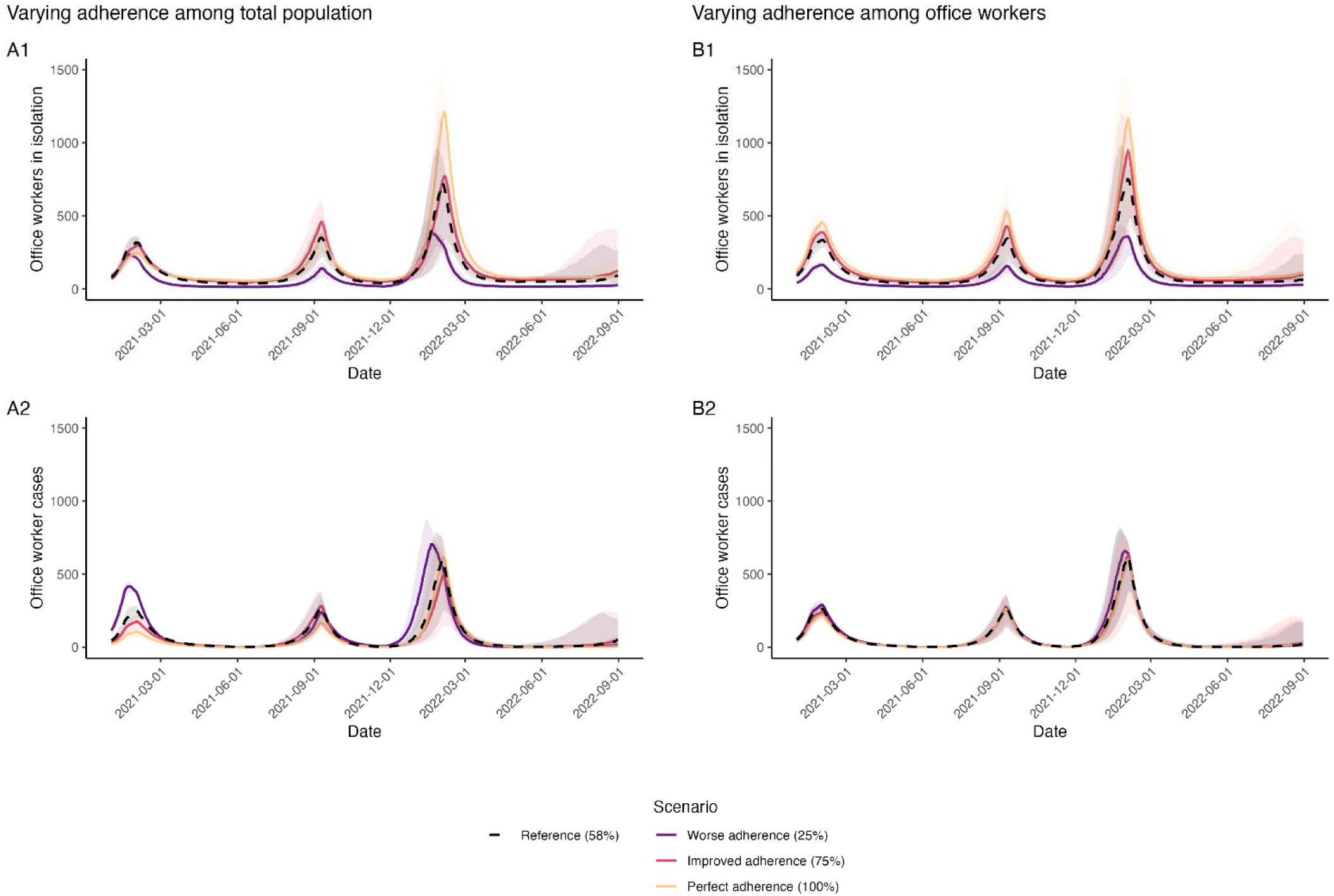
Impact of varying adherence to U.S. isolation and masking guidance on office workers isolating and office worker cases in a SARS-CoV-2 network transmission model representing Georgia, USA, from January 2021 through August 2022. Office workers in isolation includes cases who are isolating. Cases refer to symptomatic SARS-CoV-2 infections, who may or may not be isolating. Left panels (A1, A2) represent the impact of varying adherence to US CDC guidance among the total population. Right panels (B1, B2) represent the impact of varying adherence to US CDC guidance among office workers only, while others remain at reference (58%) adherence.

**Figure 6.**
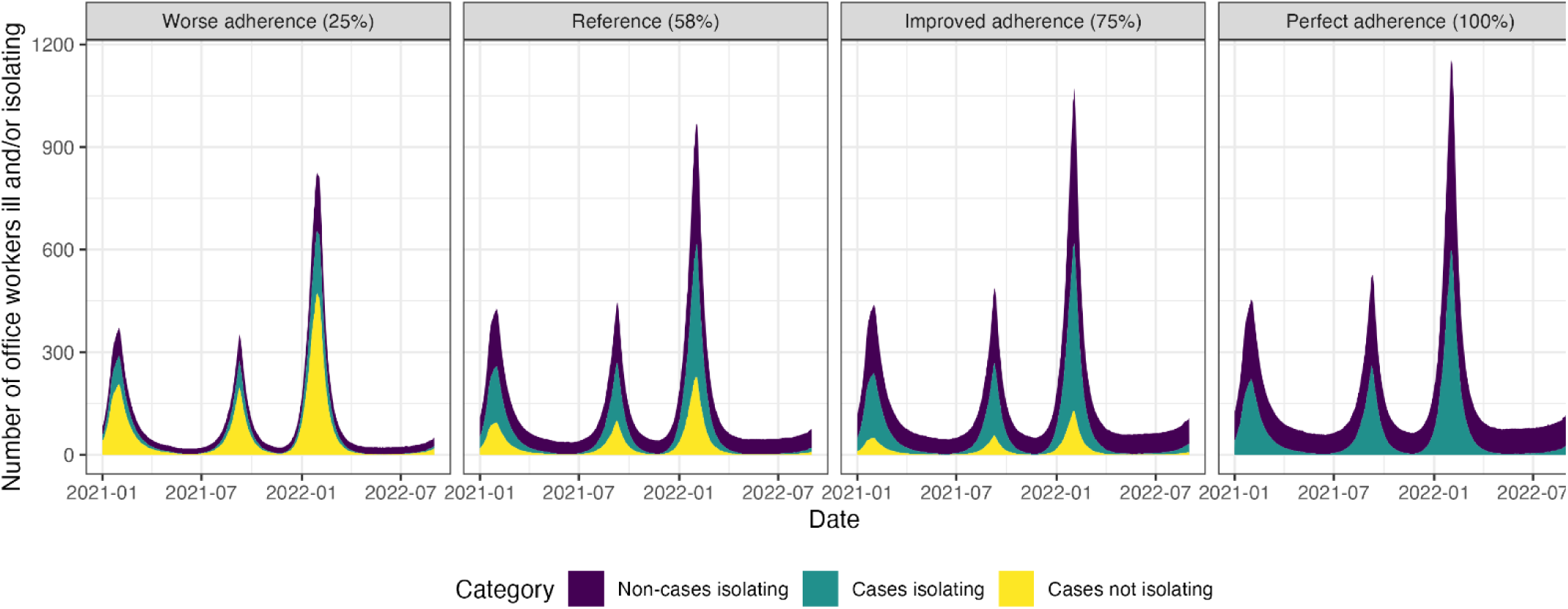
Impact of varying adherence to U.S. isolation and masking guidance among office workers on office workers isolating and office worker cases in a SARS-CoV-2 network transmission model representing Georgia, USA, from January 2021 through August 2022. Cases refer to symptomatic SARS-CoV-2 infections. Non-cases refer to all others (i.e., agents who may or may not be infected, but are not symptomatic).

Office worker outcomes were more strongly influenced by adherence to CDC guidance among the rest of the population. Improved adherence (75%) among other agents (i.e., all those not office workers) prevented 7.3% (50% SI: 2.1, 11.8) of office worker cases and 5.8% (50% SI: - 3.8, 17.4) of office worker deaths relative to the reference scenario (Table S3-S4, Figure 4).

When adherence to CDC guidance varied across the entire population, including office workers, effects were even larger; improved adherence (75%) among the total population prevented 9.3% (50% SI: 4.3, 14.5) of office worker cases and 9.6% (50% SI: -0.7, 19.7) of office worker deaths compared to the reference scenario (Figure 4). Perfect adherence (100%) among the total population had substantially greater impact, preventing over a quarter of office worker cases (28.4%; 50% SI: 23.5, 33.4) and deaths (26.9%; 50% SI: 17.2, 36.5) compared to the reference scenario (Tables S3-S4, Figure 4).

Varying the notification parameters (i.e., the probability that an office worker notified their workplace contacts after testing positive for SARS-CoV-2) had negligible impact on transmission, both among office workers and the total population, across all levels of adherence to guidance (Figure S2). On the other hand, the lower the relative contact intensity during isolation, the greater the impact on infections and deaths across levels of adherence to guidance (Figure S3, Figure S4). Furthermore, due to the impact on infections, lower levels of relative contact intensity resulted in fewer total persons entering isolation during the simulation period, even at the same level of adherence to guidance (Figure S3, Figure S4). These trends were more pronounced among the total population but were similar among office workers.

## Discussion

We used a network-based mathematical model to simulate the COVID-19 pandemic in Georgia, USA during January 2021–August 2022 and evaluated the impact of CDC isolation and masking guidance on COVID-19 among office workers in the post-vaccine period. Our calibrated model represented contact patterns at home, in the community, and in office workplaces, and reflected key dynamics in infections, hospitalizations, deaths, and vaccine coverage. We found that, in a scenario with no adherence to CDC isolation and masking guidance among office workers, there were about 27% more SARS-CoV-2 transmissions in the workplace compared to the reference scenario (58% adherence). In other words, adherence to this guidance was estimated to have prevented over a quarter of workplace-based infections. However, our model suggests improved adherence among office workers may have had marginal returns on preventing additional COVID-19 cases among office workers; improved adherence among the rest of the population appeared to be more important for preventing cases among office workers.

Although the modeled time period was after stay-at-home orders were lifted and once most businesses were once again open,^20^ an important shift towards flexible and hybrid work had occurred.^21^ This was represented in our model by using contact patterns from the Corporate Mix study,^8^ which captured social contacts among employees from five companies in Atlanta during March – June 2022. As a result, only about 12.3% of modeled SARS-CoV-2 transmissions occurred in office settings in the reference scenario, explaining the limited benefit of improved adherence to CDC isolation and masking guidance among office workers in our model.

Similarly, we found that increasing the likelihood that office workers would notify their office contacts of a positive SARS-CoV-2 test had negligible impact on transmission. This was due to a combination of 1) relatively low contacts (and transmission) in office settings; 2) lag between initial exposure, diagnosis of case, and notification of contact; and 3) imperfect prevention of further transmission due to masking. In office settings with hybrid work arrangements where social contacts are low, as was represented in our model, there may be limited benefit to enforcing isolation, masking, or contact tracing.

Since most transmission events in our model occurred outside of office settings, COVID-19 among office workers was most impacted by adherence to isolation and masking guidance by the rest of the population. The Corporate Mix study only captured social contacts for office workers, so we parametrized community contacts patterns for the rest of the population based on the widely used POLYMOD social mixing study.^14^ POLYMOD data are from pre-COVID-19 years, so they may not be representative of 2021 and 2022 contact patterns, which could bias our results. However, mobility patterns suggest that after a steep drop in March 2020, mobility returned close to pre-pandemic levels by 2021 and remained stable into 2022.^22,23^ The remaining difference in mobility could be explained by the shift to remote and hybrid work, as work-related mobility has remained lower compared to pre-pandemic.^24^ Our model suggests that community-wide adherence to isolation and masking guidance is more effective as a COVID-19 control measure than workplace-focused interventions alone.

This study has several limitations. First, as is the case with all mathematical models, our results depend on the accuracy of the contact data used to parametrize the model. Since we were focused on dynamics specific to office workers, we did not explicitly model non-office workplaces or schools, which are high-contact settings and may be important drivers of community transmission.^25^ We also only represented contacts longer than 15 minutes, which may not adequately capture all relevant contacts for SARS-CoV-2 transmission.^12^ Additionally, contact patterns likely varied significantly over the course of the modeled time period, but we were not able to represent this in our model. We believe our model appropriately captured office workplace transmission dynamics, where contacts are typically longer than 15 minutes. The greater uncertainty around non-workplace contact patterns may have affected estimates of overall community transmission but should not substantially alter our conclusions about the relative impact of adherence on office worker infections.

Second, for the feasibility and interpretability of the model, we simulated adherence to a single version of the CDC isolation and masking guidance, and adherence was set to a fixed probability in each scenario. In reality, CDC guidance changed as the pandemic evolved, and adherence likely varied over the course of the pandemic and was not uniform across guidance types (e.g., people may be more likely to isolate following a positive SARS-CoV-2 test than to mask following an exposure). Some of the effects of changing masking or isolation guidance and adherence during the study period may have been absorbed into the time-varying transmission probability modifiers used to match observed epidemic trends. Furthermore, adherence levels likely clustered within social networks, with individuals in close contact tending to either collectively follow or collectively disregard guidance. However, given the relative contact rates of office workers in vs out of the office, we believe that these nuances are unlikely to negate our qualitative finding that community-level adherence is beneficial for protecting office workers. Finally, our findings should not be generalized to other workplace settings, particularly healthcare settings such as hospitals, skilled nursing facilities, and ambulatory clinics.

In conclusion, our model suggests that moderate adherence to guidance for isolation and masking may have been important in minimizing COVID-19 cases among office workers during January 2021–August 2022. However, workplace interventions to further increase adherence to said guidance may have had limited impact when workplace contacts were low, as they were in our model, given we modeled a time period when hybrid work arrangements were common.

These results may inform future disease prevention policies in the workplace in the case of new COVID-19 variant outbreaks or if other viral respiratory diseases emerge, and highlight the public health benefits of community-wide adoption of recommended non-pharmaceutical interventions.

## Data Availability

The model code and data are available on GitHub at https://github.com/EpiModel/COVID-Corporate, and the software code is available at https://github.com/EpiModel/EpiModelCOVID/tree/Corporate-2.

https://github.com/EpiModel/EpiModelCOVID/tree/Corporate-2

## Acknowledgements

We thank the employees of the five companies that participated in the Corporate Mix study.

## CRediT Statement

**Maria Garcia Quesada:** Conceptualization, Formal analysis, Software, Visualization, Writing – original draft. **Karina Wallrafen-Sam:** Software, Writing – review and editing. **Moses Chapa:** Data curation, Writing – review and editing. **Faruque Ahmed:** Conceptualization, Writing – review and editing. **Obianuju Genevieve Aguolu:** Writing – review and editing. **Noureen Ahmed:** Writing – review and editing. **Saad B. Omer:** Conceptualization, Funding acquisition, Project administration, Writing – review and editing. **Benjamin A. Lopman:** Conceptualization, Supervision, Writing – review and editing. **Samuel M. Jenness:** Conceptualization, Methodology, Supervision, Writing – Review & Editing.

