## Supplementary Tables and Figures for "Modeling the impact of adherence to U.S. isolation and masking guidance on SARS-CoV-2 transmission in office workplaces in 2021-2022"

**Table S1.** Proportion of office workers by age group and contact patterns among office workers modeled in a in a SARS-CoV-2 network transmission model representing Georgia, USA, from January 2021 through August 2022.

| Participant Age (years) | Employed (%)[1] | Work in an office, among employed (%)[1] | Office workers (%)* | Mean daily contacts in workplace† | Within age group mixing in workplace (%) |
|---|---|---|---|---|---|
| **20-29** | 72.95 | 48.04 | 35.05 | 0.63 | 35.78 |
| **30-39** | 79.90 | 48.04 | 38.38 | 0.95 | 35.78 |
| **40-49** | 79.90 | 48.04 | 38.38 | 0.73 | 35.78 |
| **50-59** | 75.50 | 48.04 | 36.27 | 0.80 | 35.78 |
| **60-69** | 44.20 | 48.04 | 21.23 | 0.37 | 35.78 |
| **70-79** | 13.00 | 48.04 | 6.25 | NA‡ | 35.78 |
| **80-89** | 8.20 | 48.04 | 3.94 | NA‡ | 35.78 |

*Estimated as product of % employed and % of those employed who work in an office.
†Average total daily contacts across age groups were 0.72. In the model, we parameterized total daily contacts by age group for 20-69 year olds and by within age group mixing for all age groups.
‡Not available in Corporate Mix data.

1. United States Department of Labor. Employment status of the civilian noninstitutional population by age, sex, and race [Internet]. Bureau of Labor Statistics. 2024 [cited 2024 Oct 31]. Available from: https://www.bls.gov/cps/cpsaat03.htm

**Table S2.** Infection rates and infections averted overall and in the workplace at varying levels of adherence to US CDC isolation and masking guidance in a SARS-CoV-2 network transmission model representing Georgia, USA, from January 2021 through August 2022.

| Adherence to CDC guidance (%) | | Overall infections | | | Infections (transmissions) in the workplace | | |
|---|---|---|---|---|---|---|---|
| Office workers | All others | Infections per 100,000 person-days | Infections averted per 100,000 person-days | % infections averted per 100,000 person-days | Infections per 100,000 person-days | Infections averted per 100,000 person-days | % infections averted per 100,000 person-days |
| Reference (0.58) | Reference (0.58) | 131.8 (128.9, 139.0) | 0.0 (-7.2, 2.9) | 0.0 (-5.4, 2.2) | 16.2 (15.6, 16.8) | 0.0 (-0.6, 0.7) | 0.0 (-3.9, 4.1) |
| 0 | 0.58 | 140.5 (138.7, 146.5) | -8.7 (-14.7, -6.9) | -6.6 (-11.2, -5.2) | 20.6 (20.1, 21.4) | -4.4 (-5.2, -3.8) | -27.1 (-32.0, -23.7) |
| 0.25 | 0.58 | 136.5 (134.2, 149.6) | -4.7 (-17.8, -2.4) | -3.6 (-13.5, -1.8) | 18.7 (18.1, 19.7) | -2.4 (-3.5, -1.8) | -15.0 (-21.5, -11.3) |
| 0.5 | 0.58 | 132.8 (129.4, 140.1) | -1.0 (-8.3, 2.4) | -0.8 (-6.3, 1.8) | 16.8 (15.9, 17.6) | -0.6 (-1.3, 0.3) | -3.4 (-8.2, 2.1) |
| 0.75 | 0.58 | 129.6 (126.3, 145.6) | 2.2 (-13.8, 5.5) | 1.6 (-10.5, 4.2) | 15.1 (14.1, 15.9) | 1.1 (0.3, 2.1) | 7.0 (1.9, 12.9) |
| 1 | 0.58 | 126.5 (123.3, 140.1) | 5.3 (-8.3, 8.5) | 4.0 (-6.3, 6.4) | 13.4 (12.6, 14.1) | 2.9 (2.1, 3.7) | 17.7 (12.9, 22.5) |
| 0.58 | 0 | 153.0 (149.4, 159.9) | -21.2 (-28.1, -17.6) | -16.1 (-21.3, -13.4) | 16.4 (16.0, 16.9) | -0.1 (-0.6, 0.3) | -0.9 (-3.9, 1.6) |
| 0.58 | 0.25 | 144.5 (143.0, 151.9) | -12.7 (-20.1, -11.2) | -9.6 (-15.2, -8.5) | 16.5 (16.1, 17.0) | -0.3 (-0.8, 0.2) | -1.6 (-4.7, 1.0) |
| 0.58 | 0.5 | 135.3 (132.5, 146.3) | -3.5 (-14.5, -0.7) | -2.6 (-11.0, -0.5) | 16.4 (15.6, 17.2) | -0.1 (-0.9, 0.7) | -0.9 (-5.8, 4.1) |
| 0.58 | 0.75 | 123.6 (119.2, 137.4) | 8.2 (-5.6, 12.6) | 6.2 (-4.3, 9.5) | 15.7 (14.7, 16.9) | 0.5 (-0.7, 1.5) | 3.2 (-4.0, 9.2) |
| 0.58 | 1 | 108.9 (104.5, 117.1) | 22.9 (14.7, 27.3) | 17.4 (11.2, 20.7) | 14.4 (13.6, 15.6) | 1.8 (0.6, 2.6) | 11.0 (3.7, 16.2) |
| 0 | 0 | 159.0 (153.1, 166.5) | -26.2 (-33.7, -20.3) | -19.7 (-25.4, -15.3) | 19.9 (19.4, 20.9) | -3.6 (-4.6, -3.1) | -22.2 (-28.1, -19.0) |
| 0.25 | 0.25 | 147.7 (146.1, 151.8) | -14.9 (-19.0, -13.4) | -11.2 (-14.3, -10.1) | 18.6 (18.2, 19.2) | -2.3 (-2.9, -1.9) | -14.3 (-17.9, -11.5) |
| 0.5 | 0.5 | 136.6 (134.1, 145.2) | -3.8 (-12.4, -1.3) | -2.9 (-9.4, -1.0) | 16.8 (16.2, 17.7) | -0.5 (-1.4, 0.1) | -3.3 (-8.5, 0.6) |
| 0.75 | 0.75 | 121.9 (117.5, 137.3) | 10.9 (-4.5, 15.3) | 8.2 (-3.4, 11.5) | 14.5 (13.5, 15.6) | 1.8 (0.7, 2.8) | 11.1 (4.5, 17.0) |
| 1 | 1 | 100.6 (95.6, 106.4) | 32.2 (26.4, 37.2) | 24.2 (19.9, 28.0) | 11.2 (10.3, 12.2) | 5.1 (4.1, 6.0) | 31.4 (25.1, 37.0) |

To understand the impact of interventions on workplace transmission, this table differentiates infections that occur in the workplace from those that occur at home or in the community. This is different from Tables S3 and S4, which describe the overall impact of interventions on office worker cases and deaths.

**Table S3.** Symptomatic case rates and cases averted in the total population and among office workers at varying levels of adherence to US CDC isolation and masking guidance in a SARS-CoV-2 network transmission model representing Georgia, USA, from January 2021 through August 2022.

| Adherence to CDC guidance (%) | | Total population | | | Office workers | | |
|---|---|---|---|---|---|---|---|
| **Office workers** | **All others** | **Cases (ill) per 100,000 person-days** | **Number cases averted per 100,000 person-days** | **% cases averted per 100,000 person-days** | **Cases (ill) per 100,000 person-days** | **Number cases averted per 100,000 person-days** | **% cases averted per 100,000 person-days** |
| 0.58 | 0.58 | 95.5 (92.1, 98.1) | 0.0 (-2.6, 3.4) | 0.0 (-2.8, 3.6) | 101.6 (97.3, 104.6) | 0.0 (-3.0, 4.3) | 0.0 (-2.9, 4.2) |
| 0 | 0.58 | 102.2 (100.4, 104) | -6.7 (-8.5, -4.9) | -7.0 (-8.9, -5.2) | 110.9 (108.6, 113.1) | -9.3 (-11.5, -7.0) | -9.1 (-11.3, -6.9) |
| 0.25 | 0.58 | 99.2 (97, 104.2) | -3.7 (-8.7, -1.5) | -3.9 (-9.1, -1.6) | 106.9 (103.9, 111.0) | -5.3 (-9.4, -2.3) | -5.2 (-9.3, -2.2) |
| 0.5 | 0.58 | 96.1 (93.0, 98.7) | -0.6 (-3.2, 2.5) | -0.6 (-3.4, 2.6) | 102.5 (98.4, 105.8) | -0.9 (-4.2, 3.2) | -0.9 (-4.2, 3.1) |
| 0.75 | 0.58 | 94.0 (90.2, 99.1) | 1.5 (-3.6, 5.3) | 1.6 (-3.8, 5.5) | 99.6 (94.4, 104.4) | 2.0 (-2.8, 7.2) | 2.0 (-2.7, 7.1) |
| 1 | 0.58 | 90.8 (88.1, 97.6) | 4.7 (-2.1, 7.4) | 4.9 (-2.2, 7.8) | 95.4 (91.3, 101.0) | 6.2 (0.6, 10.3) | 6.1 (0.6, 10.2) |
| 0.58 | 0 | 112.2 (110.6, 115.2) | -16.7 (-19.7, -15.1) | -17.4 (-20.6, -15.9) | 120.2 (118.6, 122.7) | -18.6 (-21.1, -17) | -18.3 (-20.8, -16.7) |
| 0.58 | 0.25 | 105.7 (104.5, 107.5) | -10.2 (-12.0, -9.1) | -10.7 (-12.6, -9.5) | 113.2 (111.1, 115.0) | -11.6 (-13.4, -9.5) | -11.4 (-13.2, -9.4) |
| 0.58 | 0.5 | 98.1 (95.1, 101.5) | -2.6 (-6.0, 0.4) | -2.8 (-6.3, 0.5) | 105.0 (100.6, 108.0) | -3.4 (-6.4, 1.0) | -3.3 (-6.3, 1.0) |
| 0.58 | 0.75 | 88.8 (85.4, 94.2) | 6.7 (1.3, 10.1) | 7.0 (1.3, 10.6) | 94.2 (89.6, 99.5) | 7.5 (2.2, 12.0) | 7.3 (2.1, 11.8) |
| 0.58 | 1 | 77.6 (73.6, 82.2) | 17.9 (13.3, 21.9) | 18.7 (14.0, 23) | 82.2 (76.8, 86.7) | 19.4 (14.9, 24.9) | 19.1 (14.7, 24.5) |
| 0 | 0 | 116.9 (114.1, 120.7) | -21.3 (-25.1, -18.6) | -22.3 (-26.3, -19.4) | 126.0 (123.8, 129.9) | -24.3 (-28.1, -22.0) | -23.8 (-27.7, -21.7) |
| 0.25 | 0.25 | 108.3 (107.3, 109.7) | -12.7 (-14.2, -11.8) | -13.3 (-14.8, -12.3) | 117.0 (115.4, 118.4) | -15.3 (-16.7, -13.7) | -15.0 (-16.4, -13.4) |
| 0.5 | 0.5 | 98.9 (96.7, 101.8) | -3.3 (-6.3, -1.1) | -3.5 (-6.6, -1.2) | 105.9 (102.4, 108.8) | -4.2 (-7.1, -0.7) | -4.1 (-7.0, -0.7) |
| 0.75 | 0.75 | 87.4 (83.4, 94.0) | 8.1 (1.6, 12.2) | 8.5 (1.6, 12.7) | 92.2 (87.0, 97.3) | 9.5 (4.4, 14.7) | 9.3 (4.3, 14.5) |
| 1 | 1 | 71.0 (66.7, 75.0) | 24.5 (20.6, 28.8) | 25.7 (21.5, 30.2) | 72.8 (67.8, 77.8) | 28.9 (23.9, 33.9) | 28.4 (23.5, 33.4) |

**Table S4.** Death rates and deaths averted in the total population and among office workers at varying levels of adherence to US CDC isolation and masking guidance in a SARS-CoV-2 network transmission model representing Georgia, USA, from January 2021 through August 2022.

| **Adherence to CDC guidance (%)** | | **Total population** | | | **Office workers** | | |
|---|---|---|---|---|---|---|---|
| **Office workers** | **All others** | **Deaths per 100,000 person-days** | **Number deaths averted per 100,000 person-days** | **% deaths averted per 100,000 person-days** | **Deaths per 100,000 person-days** | **Number deaths averted per 100,000 person-days** | **% deaths averted per 100,000 person-days** |
| 0.58 | 0.58 | 0.342 (0.323, 0.362) | 0.000 (-0.021, 0.019) | 0.0 (-6.0, 5.6) | 0.386 (0.342, 0.423) | 0.000 (-0.037, 0.044) | 0.0 (-9.6, 11.5) |
| 0 | 0.58 | 0.365 (0.346, 0.384) | -0.023 (-0.042, -0.004) | -6.8 (-12.3, -1.3) | 0.409 (0.364, 0.446) | -0.023 (-0.060, 0.022) | -5.9 (-15.6, 5.7) |
| 0.25 | 0.58 | 0.357 (0.337, 0.380) | -0.015 (-0.039, 0.004) | -4.4 (-11.4, 1.3) | 0.401 (0.364, 0.445) | -0.015 (-0.059, 0.022) | -3.9 (-15.4, 5.8) |
| 0.5 | 0.58 | 0.344 (0.319, 0.363) | -0.003 (-0.021, 0.022) | -0.8 (-6.2, 6.5) | 0.379 (0.334, 0.423) | 0.007 (-0.037, 0.052) | 1.9 (-9.6, 13.4) |
| 0.75 | 0.58 | 0.337 (0.320, 0.361) | 0.004 (-0.019, 0.022) | 1.2 (-5.6, 6.4) | 0.378 (0.334, 0.416) | 0.008 (-0.030, 0.052) | 2.0 (-7.7, 13.5) |
| 1 | 0.58 | 0.330 (0.308, 0.354) | 0.012 (-0.012, 0.034) | 3.4 (-3.6, 9.9) | 0.371 (0.327, 0.416) | 0.015 (-0.030, 0.059) | 3.9 (-7.8, 15.4) |
| 0.58 | 0 | 0.382 (0.362, 0.399) | -0.041 (-0.058, -0.021) | -11.9 (-16.9, -6.1) | 0.438 (0.394, 0.476) | -0.052 (-0.089, -0.008) | -13.6 (-23.2, -2.2) |
| 0.58 | 0.25 | 0.376 (0.356, 0.394) | -0.034 (-0.052, -0.014) | -10.0 (-15.3, -4.2) | 0.423 (0.371, 0.460) | -0.037 (-0.074, 0.015) | -9.7 (-19.2, 3.8) |
| 0.58 | 0.5 | 0.351 (0.329, 0.374) | -0.010 (-0.032, 0.012) | -2.9 (-9.4, 3.6) | 0.401 (0.356, 0.446) | -0.015 (-0.060, 0.030) | -3.9 (-15.5, 7.7) |
| 0.58 | 0.75 | 0.323 (0.301, 0.342) | 0.019 (-0.001, 0.040) | 5.6 (-0.2, 11.8) | 0.364 (0.319, 0.401) | 0.023 (-0.014, 0.067) | 5.8 (-3.8, 17.4) |
| 0.58 | 1 | 0.278 (0.253, 0.293) | 0.063 (0.049, 0.088) | 18.6 (14.2, 25.8) | 0.312 (0.275, 0.349) | 0.074 (0.037, 0.111) | 19.3 (9.5, 28.9) |
| 0 | 0 | 0.387 (0.372, 0.401) | -0.041 (-0.054, -0.026) | -11.8 (-15.7, -7.4) | 0.446 (0.409, 0.483) | -0.060 (-0.098, -0.023) | -15.6 (-25.3, -6.0) |
| 0.25 | 0.25 | 0.381 (0.358, 0.399) | -0.034 (-0.052, -0.011) | -9.8 (-15.0, -3.2) | 0.430 (0.379, 0.468) | -0.045 (-0.083, 0.007) | -11.6 (-21.4, 1.7) |
| 0.5 | 0.5 | 0.351 (0.334, 0.374) | -0.004 (-0.027, 0.012) | -1.2 (-7.9, 3.6) | 0.394 (0.356, 0.438) | -0.008 (-0.052, 0.029) | -2.1 (-13.5, 7.6) |
| 0.75 | 0.75 | 0.313 (0.293, 0.337) | 0.034 (0.009, 0.054) | 9.8 (2.7, 15.5) | 0.349 (0.310, 0.388) | 0.037 (-0.003, 0.076) | 9.6 (-0.7, 19.7) |
| 1 | 1 | 0.249 (0.226, 0.268) | 0.098 (0.078, 0.121) | 28.3 (22.6, 34.9) | 0.282 (0.245, 0.319) | 0.104 (0.066, 0.141) | 26.9 (17.2, 36.5) |

**Table S5.** Modeled isolation and masking rates among the total population and office workers at varying levels of adherence to US CDC isolation and masking guidance in a SARS-CoV-2 network transmission model representing Georgia, USA, from January 2021 through August 2022.

| **Adherence to CDC guidance (%)** | | **Total population** | | **Office workers** | |
|---|---|---|---|---|---|
| **Office workers** | **All others** | **Number entering isolation per 100,000 person-days** | **Number masking due to known exposure per 100,000 person-days** | **Number entering isolation per 100,000 person-days** | **Number masking due to known exposure per 100,000 person-days** |
| Reference (0.58) | Reference (0.58) | 55.3 (53.5, 57.0) | 15.8 (15.5, 16.4) | 58.9 (56.4, 60.8) | 17.0 (16.5, 17.7) |
| 0 | 0.58 | 45.1 (44.3, 45.8) | 14.3 (14.0, 14.6) | 0.0 (0.0, 0.0) | 0.0 (0.0, 0.0) |
| 0.25 | 0.58 | 49.8 (48.7, 52.3) | 15.5 (15.2, 16.3) | 26.7 (26.0, 27.8) | 9.1 (8.8, 9.5) |
| 0.5 | 0.58 | 53.8 (52.1, 55.6) | 15.8 (15.4, 16.3) | 51.1 (49.2, 52.9) | 15.5 (15.0, 16.1) |
| 0.75 | 0.58 | 58.2 (55.9, 61.4) | 15.5 (15.2, 16.7) | 74.5 (70.8, 78.4) | 19.4 (18.8, 20.5) |
| 1 | 0.58 | 61.6 (59.5, 66.1) | 14.4 (14.1, 15.6) | 95.4 (91.3, 101) | 20.2 (19.7, 21.7) |
| 0.58 | 0 | 15.5 (15.2, 15.8) | 5.7 (5.6, 5.9) | 69.8 (68.7, 71.2) | 25.9 (25.3, 26.7) |
| 0.58 | 0.25 | 34.7 (34.3, 35.3) | 12.4 (12.1, 12.7) | 65.7 (64.5, 66.9) | 22.7 (22.2, 23.4) |
| 0.58 | 0.5 | 50.9 (49.3, 52.7) | 15.7 (15.3, 16.4) | 61.0 (58.0, 62.7) | 18.6 (18.0, 19.5) |
| 0.58 | 0.75 | 63.1 (60.6, 67.1) | 14.7 (14.3, 15.7) | 54.6 (52.1, 57.7) | 13.6 (13.1, 14.4) |
| 0.58 | 1 | 69.9 (66.5, 74.1) | 9.5 (9.1, 9.9) | 47.5 (44.6, 50.3) | 8.1 (7.7, 8.5) |
| 0 | 0 | 0.0 (0.0, 0.0) | 0.0 (0.0, 0.0) | 0.0 (0.0, 0.0) | 0.0 (0.0, 0.0) |
| 0.25 | 0.25 | 27.1 (26.8, 27.5) | 10.4 (10.2, 10.6) | 29.2 (28.7, 29.8) | 7.5 (7.2, 7.7) |
| 0.5 | 0.5 | 49.5 (48.4, 50.8) | 15.6 (15.3, 16.2) | 53.0 (51.3, 54.3) | 11.1 (10.7, 11.6) |
| 0.75 | 0.75 | 65.5 (62.6, 70.6) | 14.0 (13.6, 15.4) | 69.1 (65.3, 73.1) | 9.9 (9.4, 10.6) |
| 1 | 1 | 71.0 (66.7, 75.0) | 5.7 (5.5, 6.0) | 72.8 (67.8, 77.8) | 4.0 (3.8, 4.2) |

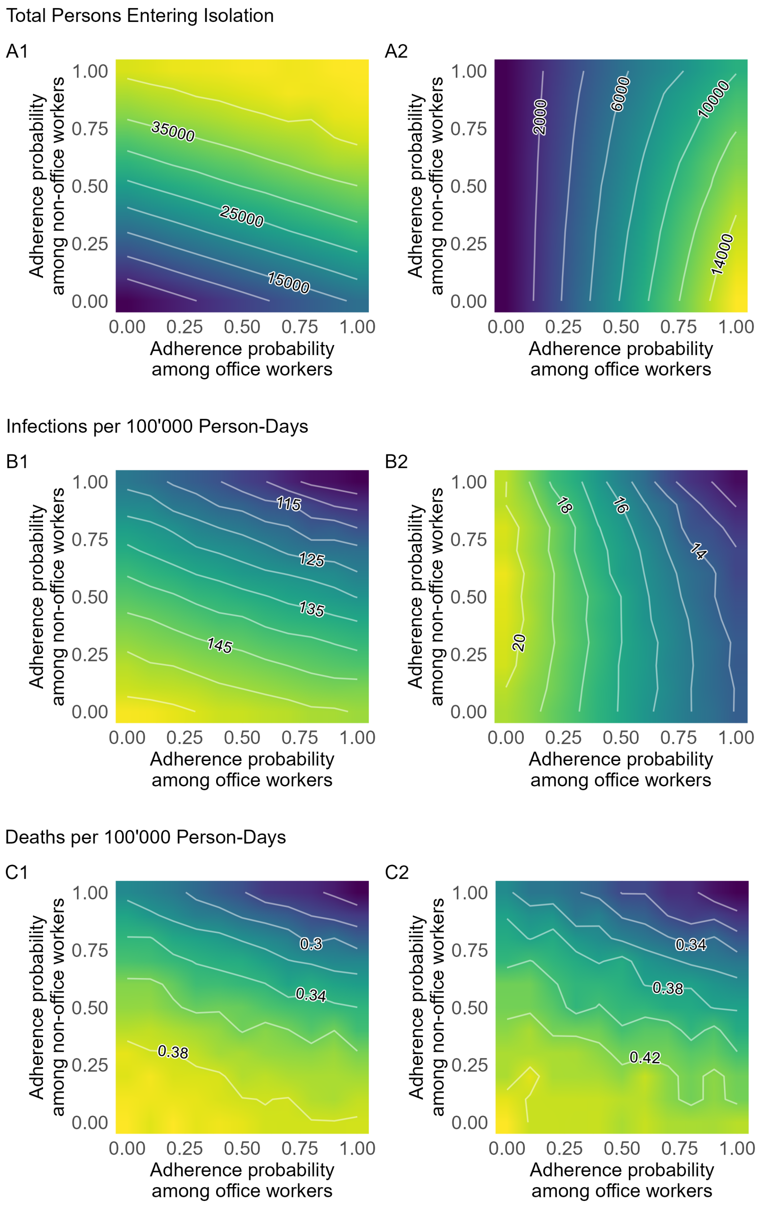


**Figure S1.** Changes in persons entering isolation, infection rates, and death rates when varying adherence to US CDC isolation and masking guidance among office workers and among all others (non-office workers) across the parameter space.

Panels on the left (A1, B1, C1) represent the impact among the total population, and panels on the right (A2, B2, and C2) represent the impact among office workers.

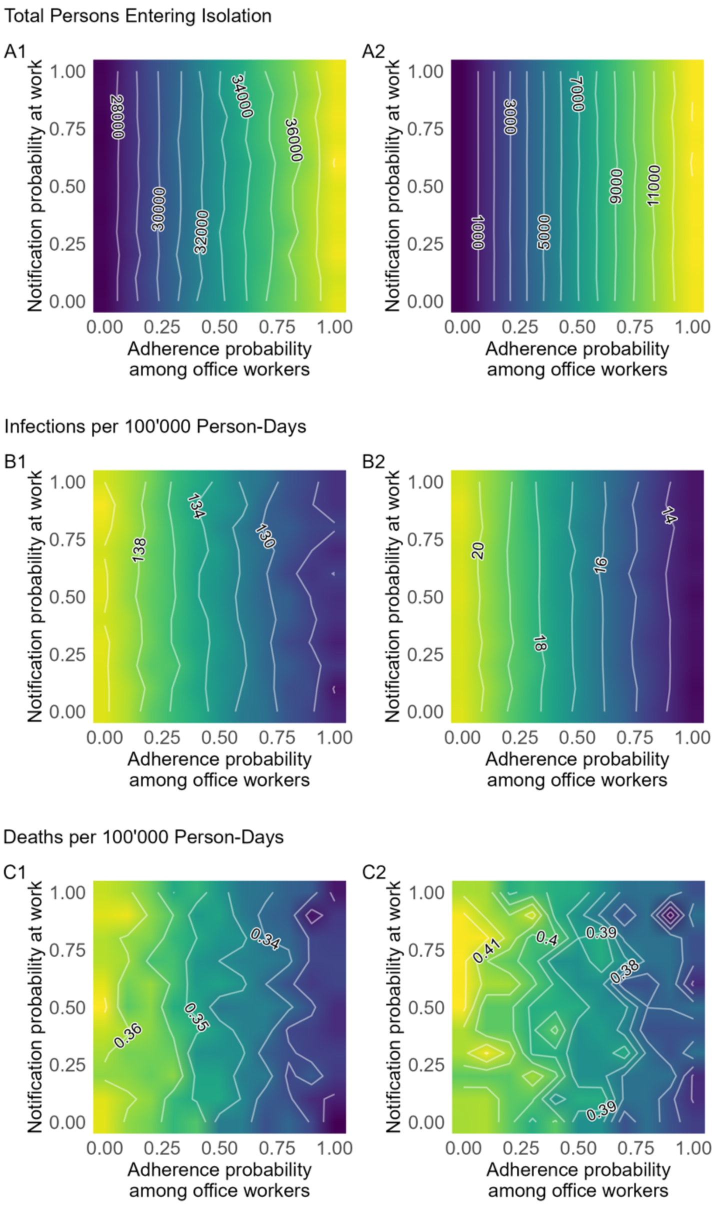


**Figure S2.** Changes in persons entering isolation, infection rates, and death rates when varying adherence to US CDC isolation and masking guidance among office workers and the probability to notify work contacts following a positive SARS-CoV-2 test across the parameter space.
Panels on the left (A1, B1, C1) represent the impact among the total population, and panels on the right (A2, B2, and C2) represent the impact among office workers.

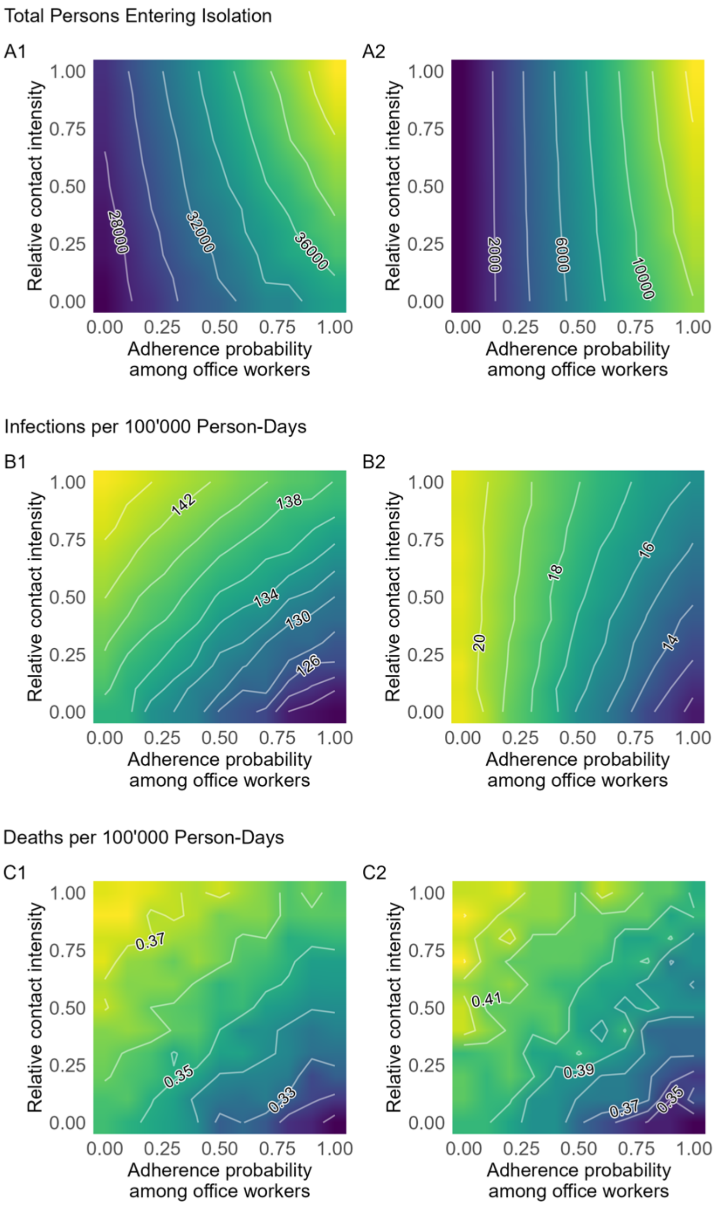


**Figure S3.** Changes in persons entering isolation, infection rates, and death rates when varying adherence to US CDC isolation and masking guidance among office workers and the relative contact intensity while isolating across the parameter space.

Panels on the left (A1, B1, C1) represent the impact among the total population, and panels on the right (A2, B2, and C2) represent the impact among office workers. Relative contact intensity is equivalent to the quality of isolation (i.e., relative contact intensity of 0.25 represents 75% reduction in contacts during isolation).

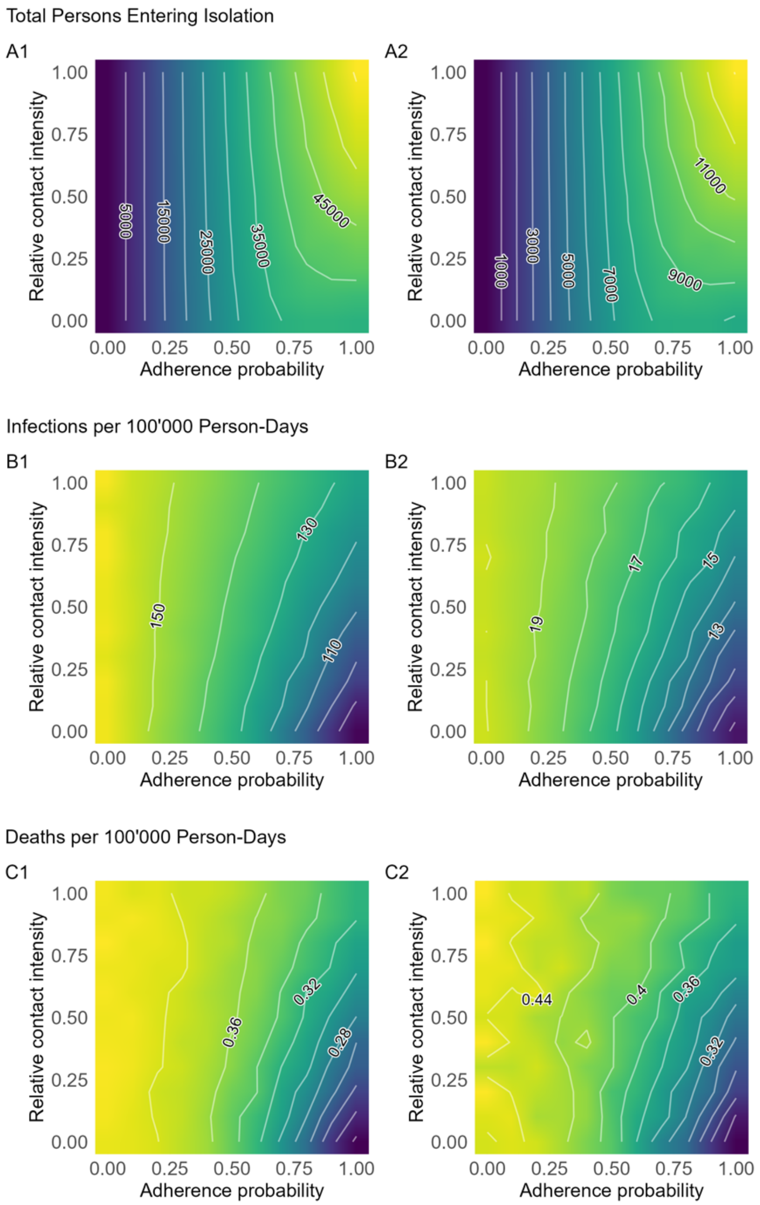


**Figure S4.** Changes in persons entering isolation, infections rates, and death rates when varying adherence to US CDC isolation and masking guidance among the total population and the relative contact intensity while isolating across the parameter space.

Panels on the left (A1, B1, C1) represent the impact among the total population, and panels on the right (A2, B2, and C2) represent the impact among office workers. Relative contact intensity is equivalent to the quality of isolation (i.e., relative contact intensity of 0.25 represents 75% reduction in contacts during isolation).

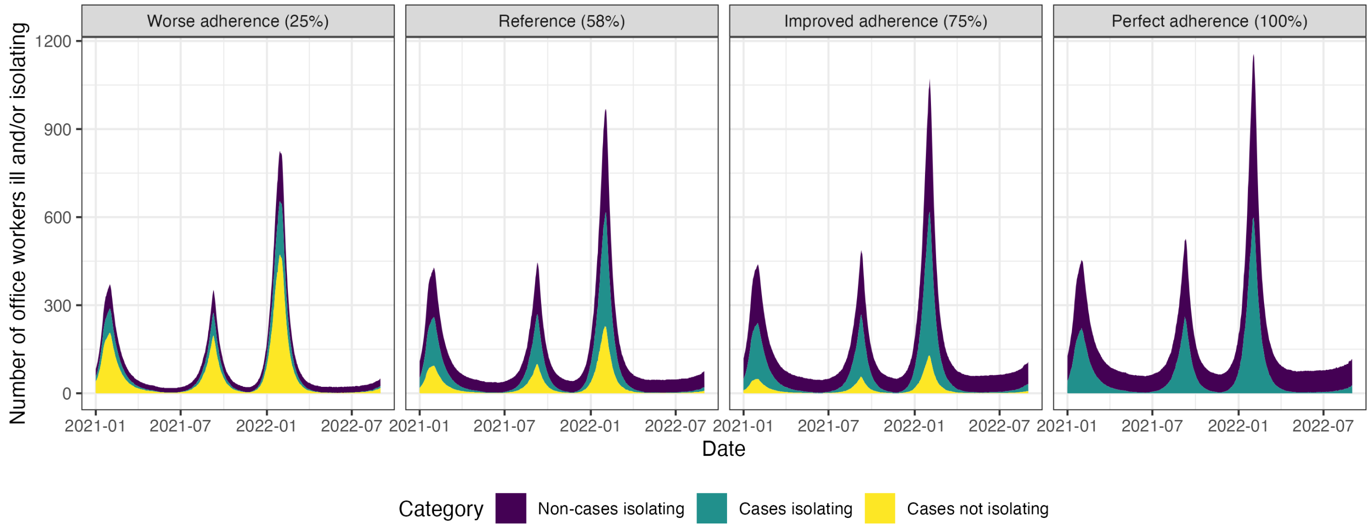


**Figure S5.** Impact of varying adherence to U.S. isolation and masking guidance among the total population on office workers isolating and office worker cases in a SARS-CoV-2 network transmission model representing Georgia, USA, from January 2021 through August 2022.
Cases refer to symptomatic SARS-CoV-2 infections. Non-cases refer to all others (i.e., agents who may or may not be infected, but are not symptomatic).
